# JOINT APPLICATION OF BRAIN MRI AND GENE EXPRESSION ATLAS TO RECONSTRUCT NMOSD PATHOPHYSIOLOGY

**DOI:** 10.1101/2023.07.19.23292876

**Authors:** Laura Cacciaguerra, Loredana Storelli, Elisabetta Pagani, Sarlota Mesaros, Vittorio Martinelli, Lucia Moiola, Marta Radaelli, Jovana Ivanovic, Olivera Tamas, Jelena Drulovic, Massimo Filippi, Maria A. Rocca

**Affiliations:** Neuroimaging Research Unit, Division of Neuroscience, IRCCS San Raffaele Scientific Institute, Milan, Italy; Neurology Unit, IRCCS San Raffaele Scientific Institute, Milan, Italy; Neurorehabilitation Unit, IRCCS San Raffaele Scientific Institute, Milan, Italy; Neurophysiology Service, IRCCS San Raffaele Scientific Institute, Milan, Italy; Vita-Salute San Raffaele University, Milan, Italy; Neurology Clinic, University Clinical Centre of Serbia, Faculty of Medicine, University of Belgrade, Belgrade, Serbia; Neurology Clinic, University Clinical Centre of Serbia, Belgrade, Serbia

**Keywords:** Neuromyelitis optica spectrum disorders, Magnetic Resonance Imaging, Gene expression.

## Abstract

**Objective:** Brain lesions in aquaporin-4-antibody-positive neuromyelitis optica spectrum disorder (AQP4+NMOSD) occur at areas of high AQP4 expression. However, the pathophysiological cascade requires additional factors such as complement. We sought to investigate the spatial association between brain damage and gene expression in AQP4+NMOSD.

**Methods:** In this multicenter cross-sectional study, we enrolled 90 patients and 94 age-matched healthy controls who underwent a 3.0/1.5 T brain MRI. In patients, brain damage was assessed through (i) T2-hyperintense lesion probability maps, (ii) white matter (WM) and grey matter (GM) atrophy on 3D T1-weighted sequences, and (iii) WM microstructural abnormalities on diffusion-tensor imaging. The association between imaging maps and the expression of 266 candidate genes in the Allen Human Brain Atlas was obtained and overrepresented biological processes were investigated with a functional-enrichment analysis.

**Results:** In AQP4+NMOSD, T2-hyperintense lesions were mainly located in the periventricular WM. GM and WM atrophy involved the visual pathway, while WM microstructural abnormalities were represented by a widespread increase of mean diffusivity. The expression of AQP4, C4a and C5 elements of complement resulted associated with all types of brain damage. Complement activation and the regulation and uptake of insulin-like growth factor were the most relevant enriched pathways. Non-specific pathways related to DNA synthesis and repair were associated with brain atrophy.

**Interpretation:** A joint application of quantitative MRI and gene expression atlas can identify in vivo the key elements of AQP4+NMOSD pathophysiology. This may pave the way to a novel type of imaging analysis helpful in understanding the pathophysiology of antibody-mediated autoimmune disorders.

## Introduction

Antibody-associated autoimmune conditions of the CNS can target surface or intracellular antigens. Intracellular autoimmunity usually represents the epiphenomenon of cytotoxic CD8+ T cell activation (e.g., in case of paraneoplastic autoimmune syndromes), while antibodies binding membrane proteins likely embody the pathogenetic element of pure autoimmune diseases with specific clinical manifestations.^1, 2^

This is the case of aquaporin-4 (AQP4) seropositive neuromyelitis optica spectrum disorder (AQP4+NMOSD), an antibody-mediated autoimmune disease targeting the AQP4 water channel, which is particularly expressed on astrocytes endfeet.^3, 4^

Pre-clinical studies demonstrated that AQP4 expression in tissues defines the susceptibility to damage of different CNS regions to AQP4+NMOSD damage. In fact, the expression of the AQP4 messenger RNA and proteins are significantly higher in the optic nerve and spinal cord,^5^ which are the main targets of the disease.^3^ Similarly, MRI studies in patients with AQP4+NMOSD demonstrated that focal brain lesions are typically located at areas with high AQP4 expression, such as the periependymal lying and the area postrema.^6^ However, this regional distribution of focal brain damage might only partially detect the heterogeneity of brain damage and pathogenesis of AQP4+NMOSD. In fact, quantitative MRI studies also documented brain atrophy, especially in the grey matter (GM),^7^ and microstructural abnormalities of the normal-appearing white matter (WM), mainly in terms of increased mean diffusivity.^8, 9^ Likewise, it is already established that tissue damage requires not only the binding between the autoantibody and AQP4, but also the activation of the complement cascade, with consequent astrocytes injury, local recruitment of granulocytes (especially eosinophils and neutrophils) and secondary demyelination and axonal loss in the the standby tissue.^10^

Recent efforts allowed the characterization of transcriptomic profiles of the human brain, leading to the definition of atlases enclosing quantitative gene expression at different brain locations, such as the Allen Human Brain Atlas (AHBA).^11^ This information can be translated to the standard reference space for MRI, the Montreal National Institute (MNI), through novel computational tools such as the Multimodal Environment for Neuroimaging and Genomic Analysis (MENGA) platform.^12^

In this study, we tried to establish whether the joint application of quantitative MRI and human brain gene expression atlases can provide an explanation to damage distribution in AQP4+NMOSD by underpinning the link with key elements of disease pathogenesis.

We investigated the spatial association between brain damage and gene expression in AQP4+NMOSD through a multimodal quantitative assessment of brain damage, including T2-hyperintense lesion distribution, WM and GM atrophy, and WM microstructural abnormalities.

In the interpretation of results, we specifically focused on immune-system elements targeted by the Food and Drug Administration (FDA)-approved treatments for AQP4+NMOSD, namely complement (eculizumab), B cells (inebilizumab), and the interleukin (IL) -6 pathway (satralizumab).

## Methods

### Ethics committee approval

Subjects’ consent was obtained according to the Declaration of Helsinki. Approval was received from the ethical standards committee of IRCCS Ospedale San Raffaele, Milan, Italy and of the University Clinical Centre of Serbia, Belgrade, Serbia, and written informed consent was obtained from all participants at the time of data acquisition.

### Participants

Brain MRI scans were acquired from 80 AQP4+NMOSD patients diagnosed according to 2015 International Panel Consensus Diagnostic criteria^3^ and 94 healthy controls (HC). AQP4-IgG serostatus was tested using a cell-based assay.^3^ Participants were recruited at two European centers: Milan (92 subjects: 40 AQP4+NMOSD and 52 HC) and Belgrade (82 subjects: 40 AQP4+NMOSD and 42 HC). Exclusion criteria were alcohol or drug abuse, history of head trauma, psychiatric comorbidities, and contraindications to MRI. On the day of the MRI acquisition, patients also underwent a neurological examination including the assessment of the Expanded Disability Status Scale (EDSS)^13^ and history collection.

### MRI acquisition protocol

Using two 3.0 T scanners (Milan: Ingenia CX and Intera Philips Medical Systems) and a 1.5 T scanner (Belgrade: Achieva Philips Medical System), the following brain sequences were collected from all subjects in a single session:

i. T2-weighted images: axial dual-echo turbo spin-echo (TSE) (Intera scanner in Milan and Belgrade) or sagittal 3D fluid attenuated inversion recovery (FLAIR) (Ingenia CX scanner in Milan);
ii. 3D T1-weighted images: axial 3D T1-weighted fast gradient-echo (Intera scanner in Milan), axial 3D T1-weighted turbo field echo (Belgrade) and sagittal 3D T1-weighted magnetization-prepared rapid gradient-echo (Ingenia scanner in Milan);
iii. Diffusion-weighted images (DWI): axial pulsed-gradient spin echo diffusion-weighted echo-planar imaging (single shell, Intera scanner in Milan and Belgrade), axial pulsed-gradient spin echo single shot diffusion-weighted echo planar imaging (3 shells, Ingenia scanner in Milan).

Details of the MRI protocols are available as Supplementary Table 1.

### MRI analysis

#### Conventional MRI analysis

Focal WM lesions were segmented on T2-weighted images using a fully-automated approach based on two 3D patch-wise convolutional neural networks^14^ (Milan, Ingenia scanner) or using a local thresholding segmentation technique (Jim 8.0 Xinapse System Ltd; Milan, Intera scanner and Belgrade) to obtain lesion masks and volumes. Head-size normalized volumes of the brain (NBV), GM (NGMV) and WM (NWMV) were measured after T1-hypointense lesion refilling^15^ using the FSL SIENAx software.^16^

#### T2-hyperintense lesion probability map in AQP4+NMOSD patients

By exploiting the transformation matrix of the voxel-based morphometry (VBM) (refer to the “Voxel-based morphometry” section for further details), we coregistered the single-subject T2-lesion masks to the standard space, obtaining a lesion probability map (i.e., 0-100% probability of a voxel of being involved by a lesion). Since only patients with brain lesions were included, and the lesion volume in the disorder is usually moderate, the lesion mask of the right hemisphere was mirrored on the left hemisphere to strength our statistical power (i.e., if corresponding voxels on the two hemispheres were involved by a lesion, the probability of having a lesion in that voxel was calculated as the sum of both probabilities). The choice of the left hemisphere was driven by the fact that only two of the six donors of the AHBA were sampled also in the right hemisphere.

#### Voxel-based morphometry

VBM analysis was used to identify regions of significant GM and WM atrophy in AQP4+NMOSD patients compared to HC. The opposite contrast was also performed as negative control.

3D T1-weighted images were segmented using the Segmentation tool available in SPM12 (i.e., GM, WM and cerebrospinal fluid) and normalized through the Diffeomorphic Anatomical Registration using Exponentiated Lie Algebra (DARTEL) registration method.^17^ GM and WM maps were modulated and smoothed (3D 8-mm Gaussian kernel). Between-group comparisons were run with a two-factor (i.e., AQP4+NMOSD and HC) and three-level (i.e., scanners) ANOVA, with age, sex, and the SIENAX-derived V-scaling as covariates. P values <0.001 were considered significant.

#### Tract-based spatial statistics (TBSS)

Differences in the microstructural composition of the whole-brain WM between AQP4+NMOSD and HC were analyzed with the TBSS (http://www.fmrib.ox.ac.uk/fsl/tbss/index.html).18 The pre-processing of images included the correction for off-resonance distortions (Milan, Ingenia CX scanner)^19^ and for movements and eddy current-induced distortions (all scanners).^19, 20^ The diffusion tensor was estimated by linear regression on diffusion-weighted imaging data at b=700/1000 s/mm^2^ (Milan, Ingenia CX scanner), b=900 s/mm^2^ (Milan, Intera scanner), or b=1000 (Belgrade). Then, maps of fractional anisotropy (FA) and mean diffusivity (MD) were derived.^21^

Individual FA images were nonlinearly registered to the FMRIB58_FA atlas within the FSL tool, and averaged to obtain a customized FA-atlas, which was thinned to obtain a WM tract “skeleton” with FA values thresholded at 0.2 to retain voxels within the WM only. Then, single-subject FA values were further projected onto the FA skeleton by searching perpendicularity to the maximum FA values.^18^

Between-group comparisons were run with a two-factor (i.e., AQP4+NMOSD and HC) and three level (i.e., scanners) ANOVA, with age and sex as covariates.

The TBSS “randomize” tool for nonparametric permutation interference was used to identify FA voxels significantly different between AQP4+NMOSD patients and HC (p<0.05 family-wise error corrected). The number of permutations was set to 5,000 and the threshold-free cluster enhancement (TFCE) option was applied. The same process was applied to study MD.

#### Spatial associations between image-derived maps and genetics

Brain-wide gene expression profiles were obtained from the AHBA, comprising gene expression data for more than 20,000 genes from about 3,700 spatially distinct tissue samples from six neurotypical adult brains.^11, 22^ We pre-selected disease-specific genes of interest using the Open Target Platform and the search term: “Neuromyelitis optica” (https://platform.opentargets.org/disease/)^23^ which were also included in the AHBA database and MENGA platform (see below), obtaining a list of 266 candidate genes (Supplementary Table 2).

To investigate the spatial association between the imaging-derived maps and the candidate gene expression, we used the MENGA platform^12^ following published guidelines.^24^ Namely, imaging maps were: the T2-hyperintense lesion probability map, the VBM-derived T-maps of GM and WM atrophy, and the FA/MD TBSS maps of microstructural WM abnormalities.

To reduce variability and ensure consistent and reproducible results^24^ the transcriptomic data from the AHBA database underwent the following steps:

i. Representative probe selection to index expression for a gene: one probe was collected as representative of a specific gene transcript for all donors. Seventy-one% of genes in the AHBA were measured with at least two probes. Thus, probe selection was performed by evaluating the distribution of the expression values: the one with the most symmetric and least skewed profile was selected to avoid the non-linearity effect on the microarray measures.^12^
ii. Normalization of the expression measure by inter-individual differences (https://github.com/BMHLab/AHBAprocessing): to address donor-specific effects and remove the inter-individual differences in expression measure each gene expression value was normalized across brain regions separately in each donor to reflect its relative expression across different brain regions. Then, Z-score normalization was obtained as follows:

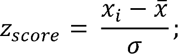

Where 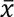 and 𝜎 are the mean and standard deviation, respectively, and 𝑥_𝑖_ is the expression value of a specific gene in a single sample.

Image maps in the MNI space were then resampled into the AHBA coordinates, separately for each donor, to obtain image-to-sample spatial correspondence according to MNI coordinates of each sample provided by the AHBA. Image data were normalized to z-scores and each image sample is estimated as the average of the voxels within a 3D window of a specified size (here set to 5 mm) centered on the MNI coordinates of the genomic sample.

Spatial association between imaging and genomic data have been therefore assessed with MENGA platform using a weighted multiple regression, with the directionality of the imaging-genomic data correlation also provided.

A measure of cross-correlation reliability is estimated with the chance likelihood, according to: ^12^

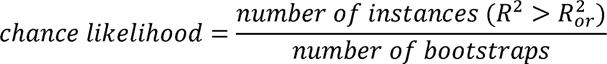

Where 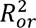 is the value of the coefficient obtained from the real data, and 𝑅^2^ is that obtained using the bootstrapped genomic data. It returns the probability that genomic data are unrelated to the image values.

Thus, smaller values of chance likelihood denoted a higher reliability of the obtained results. In this work, we set a cut-off <0.05 likelihood to assess significant spatial associations between brain damage and gene expression.

#### Enrichment analysis

To better understand functionalities, we performed a functional-enrichment analysis according to the Reactome pathway database^25^ using the g:Profiler platform (https://biit.cs.ut.ee/gprofiler/gost).^26^ Briefly, the list of the genes significantly associated with the different types of structural damage was used as input for the g:Profiler platform, which investigated overrepresented biological processes through over-representation analysis (threshold: 5-2500 genes per enriched pathway). Only significant results at Bonferroni-corrected threshold (p<0.05) were retained. The Cytoscape software version 3.9.0 was used to visualize the interactions between the enriched pathways.^27^

### Statistical analysis

Demographic, clinical, and conventional MRI variables were compared with Mann-Whitney U-test or independent samples t-test (quantitative variables, based on normality assumption) or Pearson’s Chi square test (qualitative variables).

### Data availability statement

The anonymized dataset used and analyzed during the current study is available from the corresponding author upon reasonable request.

## Results

### Demographic, clinical, and conventional MRI features of the study population

AQP4+NMOSD patients and HC had similar age, but female sex was more common in patients, as per disease epidemiology. The proportion of participants acquired on each scanner was balanced between patients and HC. The median disease duration and EDSS score in patients were 3.5 years and 4.0, respectively. Compared to HC, AQP4+NMOSD patients had whole brain and GM atrophy, but similar WM volumes. Details are reported in Table 1.

**Table 1.**
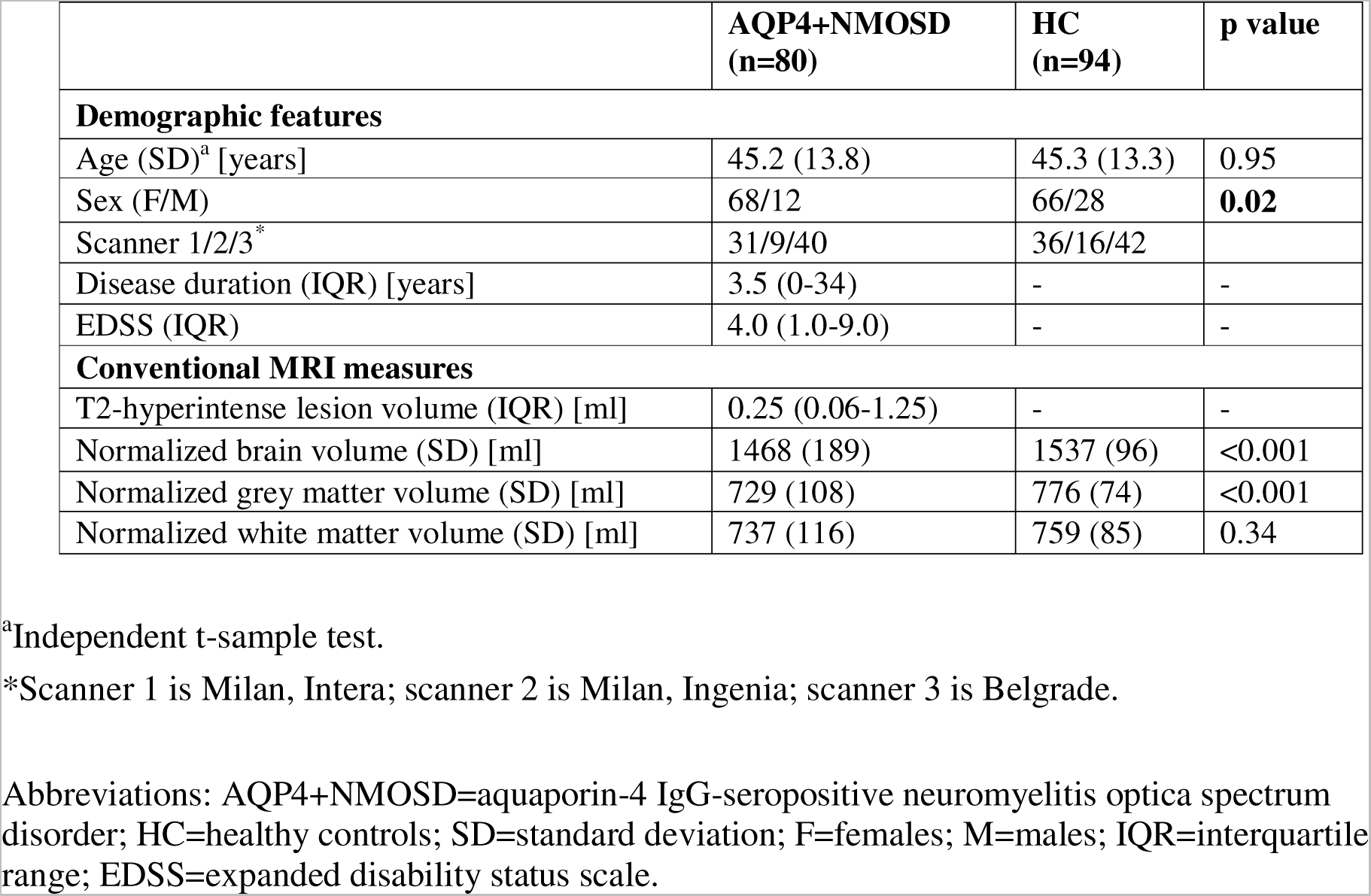
Demographic, clinical, and conventional MRI features of the study population. . Data are presented as mean (standard deviation) or median (interquartile range), according to the normality assumption. If not otherwise specified, p values refer to Mann-Whitney U-test (quantitative variables) or Pearson’s Chi square test (qualitative variables).

|  | **AQP4+NMOSD**<br>**(n=80)** | **HC**<br>**(n=94)** | **p value** |
| --- | --- | --- | --- |
| **Demographic features** |  |  |  |
| Age (SD)<sup>a</sup> [years] | 45.2 (13.8) | 45.3 (13.3) | 0.95 |
| Sex (F/M) | 68/12 | 66/28 | **0.02** |
| Scanner 1/2/3* | 31/9/40 | 36/16/42 |  |
| Disease duration (IQR) [years] | 3.5 (0-34) | - | - |
| EDSS (IQR) | 4.0 (1.0-9.0) | - | - |
| **Conventional MRI measures** |  |  |  |
| T2-hyperintense lesion volume (IQR) [ml] | 0.25 (0.06-1.25) | - | - |
| Normalized brain volume (SD) [ml] | 1468 (189) | 1537 (96) | <0.001 |
| Normalized grey matter volume (SD) [ml] | 729 (108) | 776 (74) | <0.001 |
| Normalized white matter volume (SD) [ml] | 737 (116) | 759 (85) | 0.34 |

<sup>a</sup>Independent t-sample test.
\*Scanner 1 is Milan, Intera; scanner 2 is Milan, Ingenia; scanner 3 is Belgrade.
Abbreviations: AQP4+NMOSD=aquaporin-4 IgG-seropositive neuromyelitis optica spectrum disorder; HC=healthy controls; SD=standard deviation; F=females; M=males; IQR=interquartile range; EDSS=expanded disability status scale.

### T2-hyperintense lesion probability map

Brain T2-hyperintense lesions were detected in 59/80 patients (73.7%). Lesions were mainly located in the periventricular WM. Additional locations were the subcortical WM, corpus callosum, and periaqueductal grey (Figure 1).

**Figure 1.**
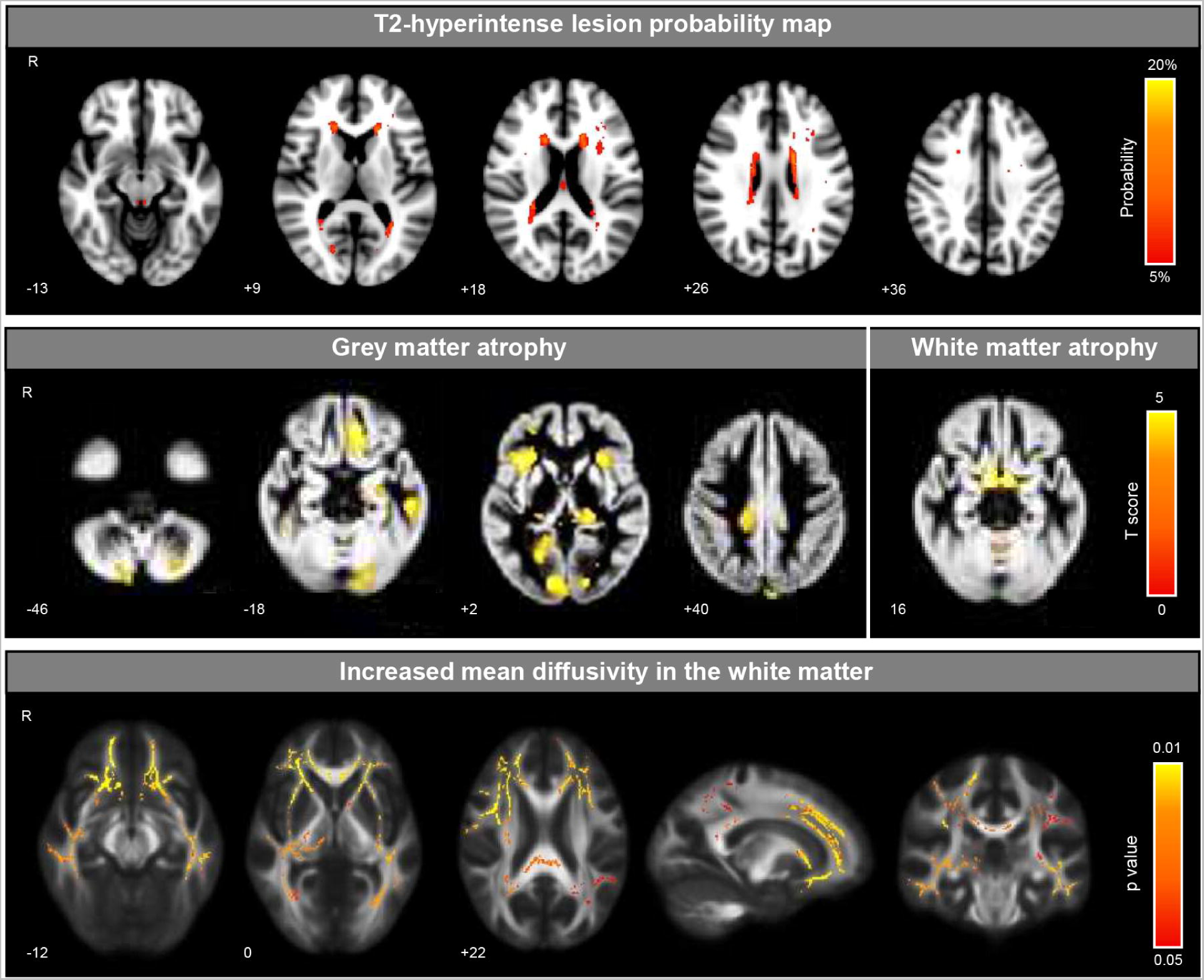
Brain damage distribution in AQP4+NMOSD patients in terms of T2-hyperintense lesion location, grey and white matter atrophy, and microstructural abnormalities. Abbreviations: R=right.

### GM and WM atrophy - VBM

Compared to HC, AQP4+NMOSD patients had significant GM atrophy in the visual cortex (left lingual gyrus, left calcarine fissure), bilateral insula, and left prefrontal cortex (orbitofrontal gyrus) (Figure 1). WM atrophy selectively involved the optic tracts, bilaterally (Figure 1). No voxels of significant atrophy were found in HC compared to AQP4+NMOSD patients.

### WM microstructural abnormalities - TBSS

Compared to HC, AQP4+NMOSD patients had increased MD in the callosal WM projecting tracts (bilateral forceps minor and major), the bilateral external capsule, posterior thalamic radiation (associative tracts to visual areas), superior longitudinal fasciculus, inferior longitudinal fasciculus (occipito-temporal associative tract), and right posterior limb of the internal capsule (corticospinal tract and medial lemniscus) (Figure 1). No significant differences in terms of FA were observed between AQP4+NMOSD patients and HC.

### Spatial association between gene expression and brain damage in AQP4+NMOSD patients

Based on results, two image-derived maps were not considered in this analysis: (i) the VBM-derived map of WM atrophy and (ii) the TBSS map of WM FA. In fact, WM atrophy selectively involved the optic tracts, which are not sampled in the AHBA. Similarly, we did not study the association between gene expression and FA in the WM since we did not find any significant difference in AQP4+NMOSD patients compared to HC (see above).

Therefore, the final spatial association between gene expression and brain damage included only the T2-hyperintense lesion probability maps, GM atrophy, and MD increase in the WM.

Significant associations are reported in Figure 2.

**Figure 2.**
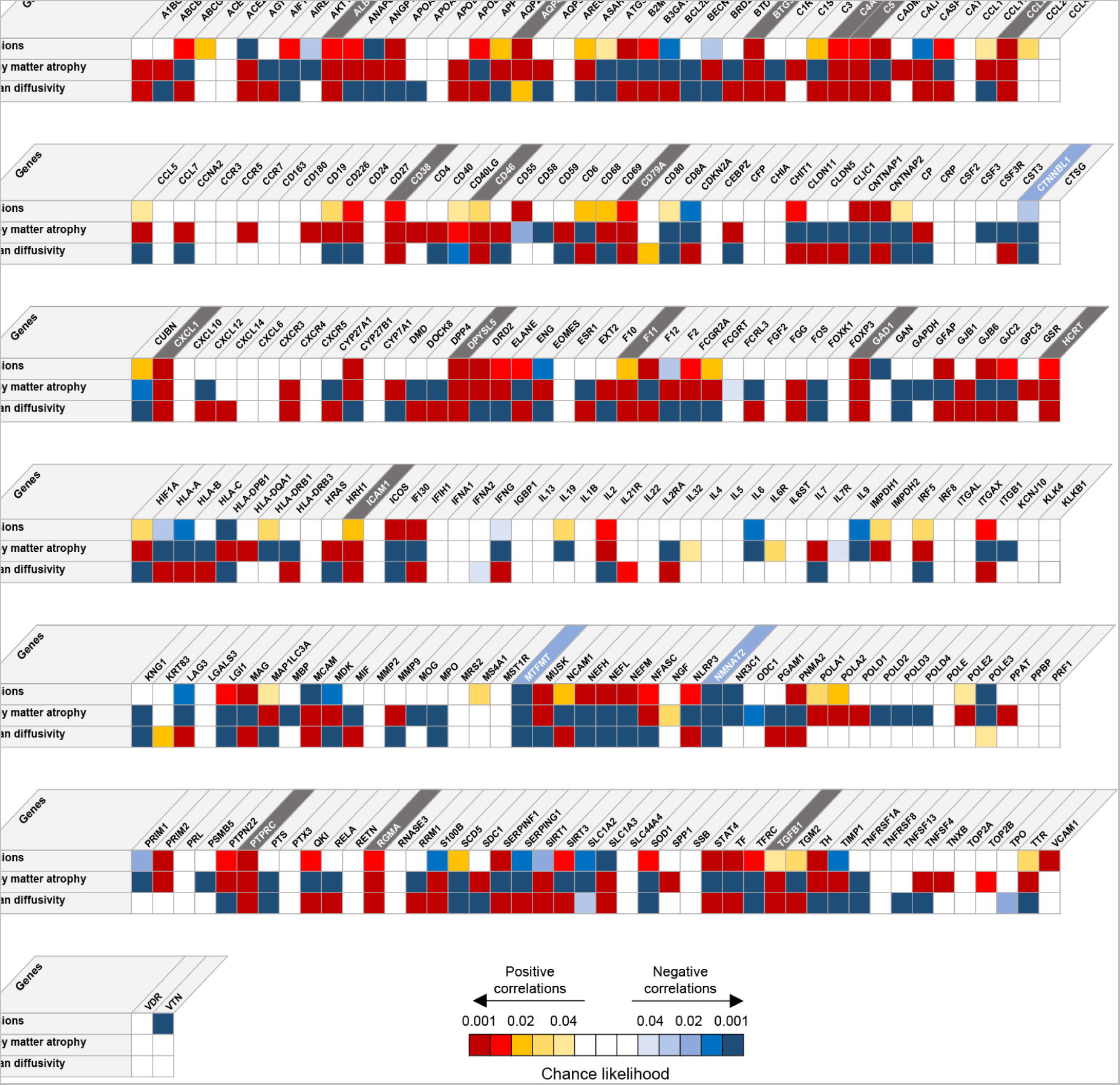
Spatial association between gene expression and brain damage in AQP4+NMOSD patients. Genes positively associated with all types of damage are highlighted in grey, while those negatively associated with all types of damage are highlighted in light blue.

The expression of AQP4 was significantly associated with all the analyzed types of damage. When we focused on targets of the FDA-approved drugs for the treatment of AQP4+NMOSD, such as eculizumab (complement inhibitor), satralizumab (IL-6 receptor inhibitor) and inebilizumab (CD19 inhibitor on B cells) we found that, likewise AQP4, a higher expression of complement factors (i.e., C5 and C4a) was associated with all types of brain damage. In contrast, IL-6 pathway showed mixed association with brain damage, since the expression of the IL-6 receptor resulted negatively associated with brain lesions and GM atrophy, but that of its transducer positively correlated with the presence of GM atrophy. The B cell marker CD19 was associated with GM atrophy only.

### Gene enrichment analysis

A total of 46 biological processes resulted enriched, of whom the association with damage was purely positive in 23, purely negative in six, and mixed positive/negative in 17. The enriched biological processes could be classified in four main clusters: pathways within (i) the immune system, (ii) mechanisms of DNA synthesis and repair, (iii) processes of peptide-ligand binding and chemokines, and (iv) regulation of the insulin-like growth factor (IGF) transport and uptake by insulin-like growth factor binding proteins (IGFBPs) (Figure 3).

**Figure 3.**
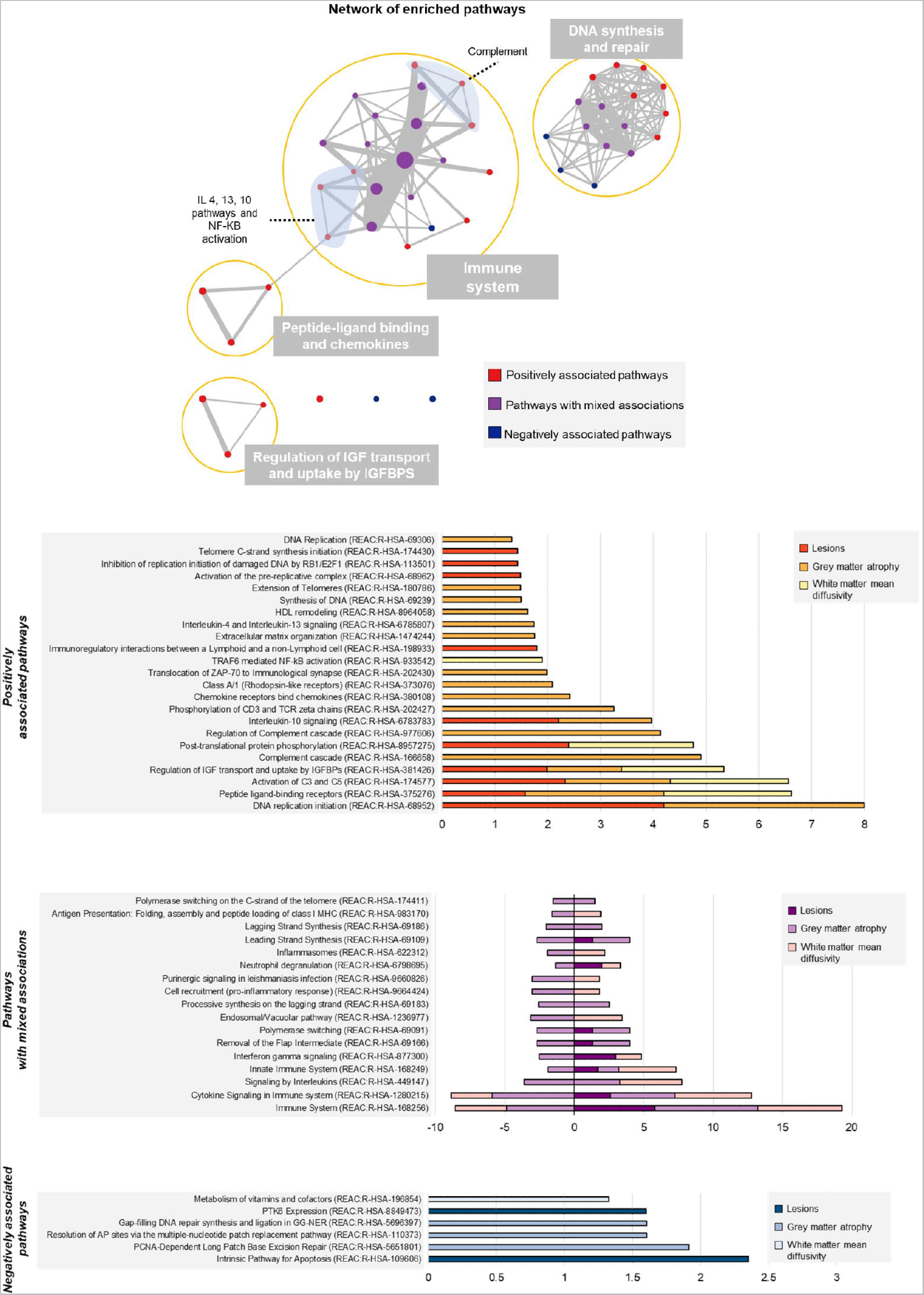
Enriched biological pathways involved in brain damage of AQP4+NMOSD. In the network nodes size represents the size of each pathway, while edges width represents the number of genes shared by the connected pathways (i.e., nodes). The barcharts show the enriched biological pathways associated with brain damage in AQP4+NMOSD. Numbers on the x axes are -log(Bonferroni-adjusted p values) of the enrichment analysis. Abbreviations: IGF=insulin-like growth factor; IGFBPS= insulin-like growth factor-binding protein; IL=interleukin; NF-KB=nuclear factor kappa-light-chain-enhancer of activated B cells; REAC=Reactome pathways database.

Complement activation, and the regulation of IGF and uptake by IGFBPs were positively associated with all types of brain damage (Figure 3). Broader processes were also significantly associated, including DNA replication initiation the response to peptide-ligand binding receptors.

Furthermore, neutrophil degranulation and the interferon gamma pathway were positively associated with the presence of lesions and increased MD in the WM. Several interleukins (IL-10, IL-4, and IL-13) were associated with the presence of GM atrophy. IL-10 also contributed to the presence of WM lesion.

Overall, the enrichment analysis disclosed a limited number of enriched biological processes spatially associated with the presence of brain lesions and increased MD in the WM. These pathways were mostly associated with AQP4+NMOSD pathophysiology.

In contrast, such an analysis identified a high number of biological processes, mainly related to DNA synthesis and repair, associated with GM atrophy (Figure 3).

## Discussion

In this study, we used a joint methodology merging quantitative measures of brain damage and the a-priori mapping of gene expression in the human brain to study its potential for the assessment and understanding of in-vivo correlates of AQP4+NMOSD pathophysiology.

Our results confirmed the key role of AQP4 expression for the development of brain lesions, atrophy, and microstructural WM abnormalities, but we were also able to identify other relevant biological pathways, namely the activation of the complement cascade and the regulation of IGF and uptake by IGFBPs, which were positively associated with all types of brain damage. IL-4, IL-13, and IL-10 signaling, neutrophil degranulation, interferon-gamma signaling, and NF-KB activation also contributed to specific types of brain injury.

The activation of the complement cascade and the recruitment of neutrophils and eosinophils at the site of damage were already identified as key steps of AQP4+NMOSD pathophysiology, as demonstrated by complement deposition and extensive granulocytes infiltrates observed in pathological tissue of patients.^10^ Therefore, the spatial association with eosinophils chemoattractants and neutrophil-related biological pathways such as IL-4 and IL-13 or interferon-gamma signaling is not surprising.

Treatments inhibiting the complement cascade, such as eculizumab and ravulizumab, were demonstrated highly effective in AQP4+NMOSD and almost invariably prevented relapses ^28, 29^. Although a head-to-head comparison was never done and studies differ for inclusion criteria and pre-enrollment disease activity, a recent metanalysis suggested that complement inhibition may be overall more effective than B cell depletion or the inhibition of the IL-6 pathway in AQP4+NMOSD.^30^

In our data, higher CD19 expression was only associated with brain GM atrophy. The same happened for IL-6 signal transducer, while other elements within the IL-6 pathway were protective or showed no association.

However, we do not believe that our results argue against the relevance of these pathways in AQP4+NMOSD pathophysiology, but rather highlight that, while the complement system exerts a local role, hence being directly involved with the presence of local brain damage, B cells and IL-6 act remotely from the periphery (e.g., by enhancing the antibody-production, favoring a pro-inflammatory environment, and increasing the blood-brain barrier permeability)^31, 32^ or in a dynamic way.

In our patients, the MRI scans were acquired during disease remission. Recent studies demonstrated that about 5% of brain lesions completely resolve and almost invariably reduce in size after the acute attack in AQP4+NMOSD.^33^ Therefore, we might have missed associations if only present during the acute phase of the disease.

We also found a significant spatial association between IL-10 signaling, lesions and GM atrophy, but literature on this topic is controversial. Both reduced and increased levels of IL-10 have been described in AQP4+NMOSD patients.^34, 35^ Despite the classical interpretation of IL-10 as an anti-inflammatory molecule, the recent SARS-CoV-2 pandemic has questioned this hypothesis since high IL-10 levels were found associated with a more severe disease course.^36^

Brain damage was also associated with the regulation of IGF transport and uptake by IGPBPs. The role of this process was not previously investigated in AQP4+NMOSD, but experimental data highlighted that IGF dysregulation (i.e., reduction) fuels autoimmunity through T reg unbalance^37^ and favors tissue damage through glutamate-induced exotoxicity^38^ and hampered remyelination.^39^ Astrocytes produce IGF in response to brain injury.^40^ Therefore, it is tempting to speculate that this protective mechanism might be deficient in AQP4+NMOSD, but future studies are warranted to confirm this result.

In general, the biological processes colocalizing with brain lesions and WM microstructural abnormalities were specific of AQP4+NMOSD pathogenesis, suggesting that lesions and WM damage are specific MRI manifestations of the disease. In contrast, GM atrophy was associated with many pathways, not always directly correlated with the pathophysiology of the disease and sometimes with unexpected directions (i.e., negative association with neutrophils degranulation and interferon-gamma signaling). In our patients, brain atrophy mainly involved structures within the visual pathways, such as the optic tracts and the occipital cortex. This suggests that atrophy is largely associated with the injury of the anterior visual pathway typically observed in the disease, leading to mechanisms of secondary neurodegeneration in posterior regions. Similar results were already obtained by others^7^ and support the lack of primary neurodegeneration in this disease. Therefore, it is likely that the complex mechanisms contributing to neurodegeneration cannot be successfully underpinned with this type of imaging analysis.

Moving to the main limitations of the study, we acknowledge the cross-sectional design and the abovementioned nuisance effect of lesion resolution. Unfortunately, biological samples of patients were unavailable at our facility, hence a biological confirmation of this approach is missing. However, our approach was able to identify complement activation as the most relevant biological process associated with brain damage in AQP4+NMOSD, which was never shown before.

Overall, we believe that the importance of this study is not to be found in the results per se, but in its translational potential. AQP4+NMOSD represents a unique research scenario where the pathogenetic element and pathophysiology are already known and proven by the efficacy of tailored treatments. Therefore, it can be used as a research model, where confirmatory results support the application of an approach to study similar disorders.

We demonstrated that a joint analysis of quantitative MRI and gene expression atlases was able to identify the main key steps of AQP4+NMOSD pathophysiology and therefore could be a tool for the understanding of antibody-associated disorders.

Nowadays, new technologies are speeding up antibody discovery,^41^ but the understanding of the underlying pathophysiology (and therefore therapeutic targets) is challenging. For instance, despite the optimization of myelin-oligodendrocyte glycoprotein (MOG)-IgG testing with a cell-based assay expressing full-length native-conformation human MOG dates back several years ago,^42^ the pathophysiology of myelin-oligodendrocyte glycoprotein antibody-associated disease is still a topic of intense scientific debate.^43, 44, 45^

Future investigations will clarify the translational power of this approach, possibly integrating results with patients’ genotyping, the measurement of serum or CSF cytokines involved in the main enriched biological processes (i.e., IL-4, IL-13, IL-10, IL-6 in this study) and biomarkers of neuronal loss.

## Acknowledgements

None.

## Author contributions

Laura Cacciaguerra: study concept, patient recruitment, clinical assessment, data analysis, statistical analysis, drafting/revising the manuscript. Loredana Storelli: data analysis, drafting/revising the manuscript. Elisabetta Pagani: data analysis, drafting/revising the manuscript. Sarlota Mesaros: patient recruitment, clinical assessment, drafting/revising the manuscript. Vittorio Martinelli: patient recruitment, clinical assessment, drafting/revising the manuscript. Lucia Moiola: patient recruitment, clinical assessment, drafting/revising the manuscript. Marta Radaelli: patient recruitment, clinical assessment, drafting/revising the manuscript. Jelena Drulovic: patient recruitment, clinical assessment, drafting/revising the manuscript. Jovana Ivanovic: patient recruitment, clinical assessment, drafting/revising the manuscript. Olivera Tamas: patient recruitment, clinical assessment, drafting/revising the manuscript. Massimo Filippi: study concept, drafting/revising the manuscript. Maria A. Rocca: study concept, drafting/revising the manuscript. All authors have approved the final version of the manuscript. Maria A. Rocca acts as guarantor and accepts full responsibility for the finished work and/or the conduct of the study, had access to the data, and controlled the decision to publish.

