## Supplementary table for "JOINT APPLICATION OF BRAIN MRI AND GENE EXPRESSION ATLAS TO RECONSTRUCT NMOSD PATHOPHYSIOLOGY"

Supplementary Table 1. Detailed description of the standardized MRI protocol at each participating center and scanner.

|  | **Scanner** | **Field strength [T]** | **Sequence** | **TR/TE [ms]** | **TI [ms]** | **b values** | **Matrix** | **Thickness**  **[mm]** |
| --- | --- | --- | --- | --- | --- | --- | --- | --- |
| **T2-w** | Milan, Intera | 3.0 | Axial dual-echo TSE | 2599/16-80 | - | - | 256x256 | 3 |
|  | Milan, Ingenia | 3.0 | Sagittal 3D FLAIR | 4800/270 | 1650 | - | 256×256 | 1 |
|  | Belgrade | 1.5 | Axial dual-echo TSE | 3141/20-100 | - | - | 256x256 | 3 |
| **T1-w** | Milan, Intera | 3.0 | Axial 3D FFE | 25/4.6 | - | - | 256x256 | 0.8 |
|  | Milan, Ingenia | 3.0 | Sagittal 3D MPRAGE | 7/3.2 | 1000 | - | 256×256 | 1 |
|  | Belgrade | 1.5 | Axial 3D TFE | 7.2/3.2 | 1000 | - | 256x256 | 1 |
| **DWI** | Milan, Intera | 3.0 | Axial pulsed-gradient spin echo diffusion-weighted echo-planar imaging | 8692/58 | - | 900 | 112×88 | 2.3 |
|  | Milan, Ingenia | 3.0 | Axial pulsed-gradient spin echo single shot diffusion-weighted echo planar imaging | 5900/78 | - | 700/1000/2855 | 112×85 | 2.3 |
|  | Belgrade | 1.5 | Axial pulsed-gradient spin echo diffusion-weighted echo-planar imaging | 6717/86 | - | 1000 | 112×110 | 2.6 |

Abbreviations: 3D=three-dimensional; DWI=diffusion-weighted images; FFE=fast field echo; FLAIR=fluid attenuated inversion recovery; mm=millimeters; MPRAGE= magnetization prepared rapid gradient echo; ms=milliseconds; T=tesla; T1-w=T1-weighted sequences; T2-w=T2-weighted sequences; TE=echo time; TFE=turbo field echo; TI=inversion time; TR=relaxation time; TSE=turbo spin-echo.

**Supplementary Table 2. List of candidate genes analyzed in the study with the overall association score (i.e., degree of association with AQP4+NMOSD according to the Open Target Platform).**

| **Ranking** | **Symbol** | **Overall association score** | **Target Name** |
| --- | --- | --- | --- |
| 1 | IL6R | 0.53257 | Interleukin 6 receptor |
| 2 | IL6ST | 0.52442 | Interleukin 6 cytokine family signal transducer |
| 3 | TOP2A | 0.47019 | DNA topoisomerase II alpha |
| 4 | IMPDH1 | 0.43426 | Inosine monophosphate dehydrogenase 1 |
| 5 | IMPDH2 | 0.43426 | Inosine monophosphate dehydrogenase 2 |
| 6 | MS4A1 | 0.40109 | Membrane spanning 4-domains A1 |
| 7 | NR3C1 | 0.40043 | Nuclear receptor subfamily 3 group C member 1 |
| 8 | CD19 | 0.39164 | CD19 molecule |
| 9 | C5 | 0.36215 | Complement C5 |
| 10 | PPAT | 0.28540 | Phosphoribosyl pyrophosphate amidotransferase |
| 11 | HLA-DQA1 | 0.13565 | Major histocompatibility complex, class II, DQ alpha 1 |
| 12 | AQP4 | 0.11965 | Aquaporin 4 |
| 13 | HLA-DRB1 | 0.09750 | Major histocompatibility complex, class II, DR beta 1 |
| 14 | IFNA1 | 0.08614 | Interferon alpha 1 |
| 15 | CSF3R | 0.08316 | Colony stimulating factor 3 receptor |
| 16 | MOG | 0.08278 | Myelin oligodendrocyte glycoprotein |
| 17 | POLA1 | 0.08223 | DNA polymerase alpha 1, catalytic subunit |
| 18 | POLA2 | 0.08223 | DNA polymerase alpha 2, accessory subunit |
| 19 | POLD1 | 0.08223 | DNA polymerase delta 1, catalytic subunit |
| 20 | POLD2 | 0.08223 | DNA polymerase delta 2, accessory subunit |
| 21 | POLD3 | 0.08223 | DNA polymerase delta 3, accessory subunit |
| 22 | POLD4 | 0.08223 | DNA polymerase delta 4, accessory subunit |
| 23 | POLE | 0.08223 | DNA polymerase epsilon, catalytic subunit |
| 24 | POLE2 | 0.08223 | DNA polymerase epsilon 2, accessory subunit |
| 25 | POLE3 | 0.08223 | DNA polymerase epsilon 3, accessory subunit |
| 26 | PRIM1 | 0.08223 | DNA primase subunit 1 |
| 27 | PRIM2 | 0.08223 | DNA primase subunit 2 |
| 28 | IL1B | 0.08036 | Interleukin 1 beta |
| 29 | SPP1 | 0.07957 | Secreted phosphoprotein 1 |
| 30 | CD38 | 0.07502 | CD38 molecule |
| 31 | APOA1 | 0.07484 | Apolipoprotein A1 |
| 32 | CD59 | 0.07430 | CD59 molecule (CD59 blood group) |
| 33 | GSR | 0.07392 | Glutathione-disulfide reductase |
| 34 | PSMB5 | 0.07392 | Proteasome 20S subunit beta 5 |
| 35 | TOP2B | 0.07392 | DNA topoisomerase II beta |
| 36 | CCR7 | 0.05802 | C-C motif chemokine receptor 7 |
| 37 | IL6 | 0.05575 | Interleukin 6 |
| 38 | TNXB | 0.05066 | Tenascin XB |
| 39 | C1S | 0.04607 | Complement c1s |
| 40 | KCNJ10 | 0.04540 | Potassium inwardly rectifying channel subfamily J member 10 |
| 41 | CD58 | 0.04396 | CD58 molecule |
| 42 | GFAP | 0.04074 | Glial fibrillary acidic protein |
| 43 | FCRL3 | 0.04071 | Fc receptor like 3 |
| 44 | CXCL10 | 0.04054 | C-X-C motif chemokine ligand 10 |
| 45 | IL7 | 0.03909 | Interleukin 7 |
| 46 | F11 | 0.03696 | Coagulation factor XI |
| 47 | F12 | 0.03696 | Coagulation factor XII |
| 48 | HRH1 | 0.03696 | Histamine receptor H1 |
| 49 | KLKB1 | 0.03696 | Kallikrein B1 |
| 50 | MBP | 0.03688 | Myelin basic protein |
| 51 | RRM1 | 0.03326 | Ribonucleotide reductase catalytic subunit M1 |
| 52 | AIRE | 0.03253 | Autoimmune regulator |
| 53 | ICOS | 0.03104 | Inducible T cell costimulator |
| 54 | NMNAT2 | 0.03088 | Nicotinamide nucleotide adenylyltransferase 2 |
| 55 | CLIC1 | 0.03085 | Chloride intracellular channel 1 |
| 56 | GJB1 | 0.02938 | Gap junction protein beta 1 |
| 57 | CSF3 | 0.02801 | Colony stimulating factor 3 |
| 58 | IL7R | 0.02716 | Interleukin 7 receptor |
| 59 | RETN | 0.02716 | Resistin |
| 60 | CD40LG | 0.02632 | CD40 ligand |
| 61 | SDC1 | 0.02605 | Syndecan 1 |
| 62 | IL4 | 0.02520 | Interleukin 4 |
| 63 | MMP9 | 0.02493 | Matrix metallopeptidase 9 |
| 64 | SLC44A4 | 0.02455 | Solute carrier family 44 member 4 |
| 65 | DPYSL5 | 0.02370 | Dihydropyrimidinase like 5 |
| 66 | ALB | 0.02294 | Albumin |
| 67 | TGM2 | 0.02217 | Transglutaminase 2 |
| 68 | GJC2 | 0.02215 | Gap junction protein gamma 2 |
| 69 | CXCR3 | 0.02204 | C-X-C motif chemokine receptor 3 |
| 70 | S100B | 0.02191 | S100 calcium binding protein B |
| 71 | HLA-DPB1 | 0.02147 | Major histocompatibility complex, class II, DP beta 1 |
| 72 | CD55 | 0.02096 | CD55 molecule (Cromer blood group) |
| 73 | IFIH1 | 0.02070 | Interferon induced with helicase C domain 1 |
| 74 | GAN | 0.01982 | Gigaxonin |
| 75 | CD8A | 0.01892 | CD8a molecule |
| 76 | TH | 0.01891 | Tyrosine hydroxylase |
| 77 | SLC1A2 | 0.01870 | Solute carrier family 1 member 2 |
| 78 | TIMP1 | 0.01848 | TIMP metallopeptidase inhibitor 1 |
| 79 | IFI30 | 0.01846 | IFI30 lysosomal thiol reductase |
| 80 | CRP | 0.01762 | C-reactive protein |
| 81 | IRF5 | 0.01682 | Interferon regulatory factor 5 |
| 82 | CD4 | 0.01628 | CD4 molecule |
| 83 | AKT1 | 0.01626 | AKT serine/threonine kinase 1 |
| 84 | ANGPT1 | 0.01626 | Angiopoietin 1 |
| 85 | QKI | 0.01626 | QKI, KH domain containing RNA binding |
| 86 | CD226 | 0.01608 | CD226 molecule |
| 87 | RELA | 0.01571 | RELA proto-oncogene, NF-kb subunit |
| 88 | C4A | 0.01524 | Complement C4A (Rodgers blood group) |
| 89 | NEFL | 0.01515 | Neurofilament light chain |
| 90 | BECN1 | 0.01478 | Beclin 1 |
| 91 | CCL7 | 0.01478 | C-C motif chemokine ligand 7 |
| 92 | MIF | 0.01404 | Macrophage migration inhibitory factor |
| 93 | IL22 | 0.01384 | Interleukin 22 |
| 94 | CCL4 | 0.01330 | C-C motif chemokine ligand 4 |
| 95 | IL32 | 0.01277 | Interleukin 32 |
| 96 | NGF | 0.01263 | Nerve growth factor |
| 97 | IL2RA | 0.01238 | Interleukin 2 receptor subunit alpha |
| 98 | AQP5 | 0.01222 | Aquaporin 5 |
| 99 | CYP27B1 | 0.01222 | Cytochrome P450 family 27 subfamily B member 1 |
| 100 | NEFH | 0.01218 | Neurofilament heavy chain |
| 101 | CYP7A1 | 0.01202 | Cytochrome P450 family 7 subfamily A member 1 |
| 102 | MMP2 | 0.01186 | Matrix metallopeptidase 2 |
| 103 | APOE | 0.01183 | Apolipoprotein E |
| 104 | ICAM1 | 0.01177 | Intercellular adhesion molecule 1 |
| 105 | HLA-DRB3 | 0.01146 | Major histocompatibility complex, class II, DR beta 3 |
| 106 | CSF2 | 0.01134 | Colony stimulating factor 2 |
| 107 | IFNG | 0.01133 | Interferon gamma |
| 108 | ABCG2 | 0.01109 | ATP binding cassette subfamily G member 2 (Junior blood group) |
| 109 | CXCL1 | 0.01109 | C-X-C motif chemokine ligand 1 |
| 110 | PPBP | 0.01109 | Pro-platelet basic protein |
| 111 | VCAM1 | 0.01056 | Vascular cell adhesion molecule 1 |
| 112 | APOA4 | 0.01035 | Apolipoprotein A4 |
| 113 | CXCL6 | 0.01035 | C-X-C motif chemokine ligand 6 |
| 114 | ESR1 | 0.01035 | Estrogen receptor 1 |
| 115 | MAP1LC3A | 0.01035 | Microtubule associated protein 1 light chain 3 alpha |
| 116 | IGBP1 | 0.01031 | Immunoglobulin binding protein 1 |
| 117 | APOB | 0.00961 | Apolipoprotein B |
| 118 | A1BG | 0.00924 | Alpha-1-B glycoprotein |
| 119 | TGFB1 | 0.00902 | Transforming growth factor beta 1 |
| 120 | SOD1 | 0.00887 | Superoxide dismutase 1 |
| 121 | TPO | 0.00887 | Thyroid peroxidase |
| 122 | PTPN22 | 0.00861 | Protein tyrosine phosphatase non-receptor type 22 |
| 123 | GJB6 | 0.00856 | Gap junction protein beta 6 |
| 124 | CALR | 0.00848 | Calreticulin |
| 125 | IL2 | 0.00847 | Interleukin 2 |
| 126 | AIF1 | 0.00832 | Allograft inflammatory factor 1 |
| 127 | CASP3 | 0.00832 | Caspase 3 |
| 128 | PRF1 | 0.00832 | Perforin 1 |
| 129 | CXCL12 | 0.00828 | C-X-C motif chemokine ligand 12 |
| 130 | CXCL14 | 0.00819 | C-X-C motif chemokine ligand 14 |
| 131 | AGT | 0.00813 | Angiotensinogen |
| 132 | CAT | 0.00813 | Catalase |
| 133 | C3 | 0.00796 | Complement C3 |
| 134 | CD79A | 0.00795 | CD79a molecule |
| 135 | BTD | 0.00739 | Biotinidase |
| 136 | DOCK8 | 0.00739 | Dedicator of cytokinesis 8 |
| 137 | DPP4 | 0.00739 | Dipeptidyl peptidase 4 |
| 138 | FGG | 0.00739 | Fibrinogen gamma chain |
| 139 | LAG3 | 0.00739 | Lymphocyte activating 3 |
| 140 | SERPINF1 | 0.00739 | Serpin family F member 1 |
| 141 | TNFRSF8 | 0.00739 | TNF receptor superfamily member 8 |
| 142 | SSB | 0.00739 | Small RNA binding exonuclease protection factor La |
| 143 | ITGAL | 0.00665 | Integrin subunit alpha L |
| 144 | BTG3 | 0.00615 | BTG anti-proliferation factor 3 |
| 145 | TNFSF13 | 0.00607 | TNF superfamily member 13 |
| 146 | CCL11 | 0.00591 | C-C motif chemokine ligand 11 |
| 147 | CCL17 | 0.00591 | C-C motif chemokine ligand 17 |
| 148 | CCR5 | 0.00591 | C-C motif chemokine receptor 5 |
| 149 | IL9 | 0.00591 | Interleukin 9 |
| 150 | CD163 | 0.00561 | CD163 molecule |
| 151 | BCL2L1 | 0.00554 | BCL2 like 1 |
| 152 | F2 | 0.00524 | Coagulation factor II, thrombin |
| 153 | AREG | 0.00517 | Amphiregulin |
| 154 | CCNA2 | 0.00517 | Cyclin A2 |
| 155 | SIRT1 | 0.00517 | Sirtuin 1 |
| 156 | LGI1 | 0.00499 | Leucine rich glioma inactivated 1 |
| 157 | PRL | 0.00480 | Prolactin |
| 158 | TNFRSF1A | 0.00480 | TNF receptor superfamily member 1A |
| 159 | HLA-C | 0.00478 | Major histocompatibility complex, class I, C |
| 160 | PTPRC | 0.00462 | Protein tyrosine phosphatase receptor type C |
| 161 | BRD2 | 0.00460 | Bromodomain containing 2 |
| 162 | NFASC | 0.00451 | Neurofascin |
| 163 | CD24 | 0.00443 | CD24 molecule |
| 164 | CD6 | 0.00443 | CD6 molecule |
| 165 | CHIA | 0.00443 | Chitinase acidic |
| 166 | CST3 | 0.00443 | Cystatin C |
| 167 | GAPDH | 0.00443 | Glyceraldehyde-3-phosphate dehydrogenase |
| 168 | IRF8 | 0.00443 | Interferon regulatory factor 8 |
| 169 | PGAM1 | 0.00443 | Phosphoglycerate mutase 1 |
| 170 | PTS | 0.00443 | 6-pyruvoyltetrahydropterin synthase |
| 171 | SCD5 | 0.00443 | Stearoyl-coa desaturase 5 |
| 172 | VDR | 0.00443 | Vitamin D receptor |
| 173 | CD27 | 0.00432 | CD27 molecule |
| 174 | MAG | 0.00425 | Myelin associated glycoprotein |
| 175 | ATG5 | 0.00407 | Autophagy related 5 |
| 176 | CCL2 | 0.00386 | C-C motif chemokine ligand 2 |
| 177 | ACE | 0.00370 | Angiotensin I converting enzyme |
| 178 | ACE2 | 0.00370 | Angiotensin converting enzyme 2 |
| 179 | ANAPC1 | 0.00370 | Anaphase promoting complex subunit 1 |
| 180 | ASAH1 | 0.00370 | N-acylsphingosine amidohydrolase 1 |
| 181 | B2M | 0.00370 | Beta-2-microglobulin |
| 182 | C1R | 0.00370 | Complement c1r |
| 183 | CD40 | 0.00370 | CD40 molecule |
| 184 | CD80 | 0.00370 | CD80 molecule |
| 185 | CLDN11 | 0.00370 | Claudin 11 |
| 186 | CTNNBL1 | 0.00370 | Catenin beta like 1 |
| 187 | CXCR4 | 0.00370 | C-X-C motif chemokine receptor 4 |
| 188 | CXCR5 | 0.00370 | C-X-C motif chemokine receptor 5 |
| 189 | DMD | 0.00370 | Dystrophin |
| 190 | ENG | 0.00370 | Endoglin |
| 191 | EOMES | 0.00370 | Eomesodermin |
| 192 | EXT2 | 0.00370 | Exostosin glycosyltransferase 2 |
| 193 | FOS | 0.00370 | Fos proto-oncogene, AP-1 transcription factor subunit |
| 194 | FOXK1 | 0.00370 | Forkhead box K1 |
| 195 | GPC5 | 0.00370 | Glypican 5 |
| 196 | HIF1A | 0.00370 | Hypoxia inducible factor 1 subunit alpha |
| 197 | HRAS | 0.00370 | Hras proto-oncogene, gtpase |
| 198 | IFNA2 | 0.00370 | Interferon alpha 2 |
| 199 | KRT83 | 0.00370 | Keratin 83 |
| 200 | MDK | 0.00370 | Midkine |
| 201 | MST1R | 0.00370 | Macrophage stimulating 1 receptor |
| 202 | MTFMT | 0.00370 | Mitochondrial methionyl-trna formyltransferase |
| 203 | NEFM | 0.00370 | Neurofilament medium chain |
| 204 | SIRT3 | 0.00370 | Sirtuin 3 |
| 205 | TFRC | 0.00370 | Transferrin receptor |
| 206 | TTR | 0.00370 | Transthyretin |
| 207 | ELANE | 0.00368 | Elastase, neutrophil expressed |
| 208 | IL5 | 0.00351 | Interleukin 5 |
| 209 | VTN | 0.00351 | Vitronectin |
| 210 | TF | 0.00333 | Transferrin |
| 211 | PTX3 | 0.00316 | Pentraxin 3 |
| 212 | NLRP3 | 0.00311 | NLR family pyrin domain containing 3 |
| 213 | CCL5 | 0.00296 | C-C motif chemokine ligand 5 |
| 214 | CD180 | 0.00296 | CD180 molecule |
| 215 | CUBN | 0.00296 | Cubilin |
| 216 | CYP27A1 | 0.00296 | Cytochrome P450 family 27 subfamily A member 1 |
| 217 | FOXP3 | 0.00296 | Forkhead box P3 |
| 218 | ITGB1 | 0.00296 | Integrin subunit beta 1 |
| 219 | KLK4 | 0.00296 | Kallikrein related peptidase 4 |
| 220 | KNG1 | 0.00296 | Kininogen 1 |
| 221 | SLC1A3 | 0.00296 | Solute carrier family 1 member 3 |
| 222 | RGMA | 0.00294 | Repulsive guidance molecule BMP co-receptor a |
| 223 | ABCB6 | 0.00259 | ATP binding cassette subfamily B member 6 (Langereis blood group) |
| 224 | CD46 | 0.00259 | CD46 molecule |
| 225 | CHIT1 | 0.00259 | Chitinase 1 |
| 226 | IL13 | 0.00259 | Interleukin 13 |
| 227 | CD68 | 0.00222 | CD68 molecule |
| 228 | CD69 | 0.00222 | CD69 molecule |
| 229 | CFP | 0.00222 | Complement factor properdin |
| 230 | CNTNAP2 | 0.00222 | Contactin associated protein 2 |
| 231 | CP | 0.00222 | Ceruloplasmin |
| 232 | F10 | 0.00222 | Coagulation factor X |
| 233 | FCGRT | 0.00222 | Fc gamma receptor and transporter |
| 234 | LGALS3 | 0.00222 | Galectin 3 |
| 235 | MUSK | 0.00222 | Muscle associated receptor tyrosine kinase |
| 236 | NCAM1 | 0.00222 | Neural cell adhesion molecule 1 |
| 237 | ODC1 | 0.00222 | Ornithine decarboxylase 1 |
| 238 | PNMA2 | 0.00222 | PNMA family member 2 |
| 239 | RNASE3 | 0.00222 | Ribonuclease A family member 3 |
| 240 | SERPING1 | 0.00222 | Serpin family G member 1 |
| 241 | APP | 0.00201 | Amyloid beta precursor protein |
| 242 | AQP2 | 0.00185 | Aquaporin 2 |
| 243 | CCL24 | 0.00185 | C-C motif chemokine ligand 24 |
| 244 | HCRT | 0.00185 | Hypocretin neuropeptide precursor |
| 245 | HLA-B | 0.00185 | Major histocompatibility complex, class I, B |
| 246 | ITGAX | 0.00185 | Integrin subunit alpha X |
| 247 | B3GAT1 | 0.00148 | Beta-1,3-glucuronyltransferase 1 |
| 248 | CADM4 | 0.00148 | Cell adhesion molecule 4 |
| 249 | CCR3 | 0.00148 | C-C motif chemokine receptor 3 |
| 250 | CDKN2A | 0.00148 | Cyclin dependent kinase inhibitor 2A |
| 251 | CEBPZ | 0.00148 | CCAAT enhancer binding protein zeta |
| 252 | CLDN5 | 0.00148 | Claudin 5 |
| 253 | CNTNAP1 | 0.00148 | Contactin associated protein 1 |
| 254 | CTSG | 0.00148 | Cathepsin G |
| 255 | DRD2 | 0.00148 | Dopamine receptor D2 |
| 256 | FCGR2A | 0.00148 | Fc gamma receptor iia |
| 257 | FGF2 | 0.00148 | Fibroblast growth factor 2 |
| 258 | GAD1 | 0.00148 | Glutamate decarboxylase 1 |
| 259 | HLA-A | 0.00148 | Major histocompatibility complex, class I, A |
| 260 | IL19 | 0.00148 | Interleukin 19 |
| 261 | IL21R | 0.00148 | Interleukin 21 receptor |
| 262 | MCAM | 0.00148 | Melanoma cell adhesion molecule |
| 263 | MPO | 0.00148 | Myeloperoxidase |
| 264 | MRS2 | 0.00148 | Magnesium transporter MRS2 |
| 265 | STAT4 | 0.00148 | Signal transducer and activator of transcription 4 |
| 266 | TNFSF4 | 0.00148 | TNF superfamily member 4 |
